# A Unified Multi-State Approach for Investigating the Dynamics of Chronic and Infectious Diseases

**DOI:** 10.64898/2026.01.17.26344210

**Authors:** Ming Ding

**Affiliations:** Department of Emergency Medicine, School of Medicine, University of North Carolina at Chapel Hill, Chapel Hill, NC, USA; Department of Epidemiology, Gillings School of Global Public Health, University of North Carolina at Chapel Hill, Chapel Hill, NC, USA

## Abstract

Infectious diseases and chronic diseases are two major fields in epidemiology that have traditionally been studied separately because of their distinct etiologies and modeling methods. Infectious disease data are typically collected at an aggregated level and analyzed using compartmental models, most commonly the susceptible (S), infectious (I), and recovered (R) (SIR) model, whereas chronic disease data are usually collected at the individual level and analyzed using multi-state survival models. Previous studies have pointed out the link between compartmental models and survival analysis by reconstructing the aggregated infection disease data into individual-level data. However, these studies have largely focused on the two-state transition from S to I state, and few studies have simultaneously modeled the three-state process, S, I, and R. In this paper, we propose to use a discrete-time multi-state framework to model the three-state progression of infectious disease. We first introduce and compare the underlying methodological foundations for modeling infectious disease and chronic disease dynamics, then show the link between compartment models and multi-state models, and finally present how infectious disease can be modeled using the multi-state framework under the two scenarios: 1) all S, I, and R states are observed, and 2) only the I state is observed, with the R state treated as latent. In the application, we applied the multi-state approach to estimate the dynamics of influenza using the data in a British boarding school in 1978, where only the infected cases were observed over time. The estimated recovery rate was 0.42 and the corresponding contact rate was0.91 (95% CI: 0.84, 0.98). The basic reproductive number was 2.17 (95% CI: 2.00, 2.33), which declined to approximately 1 by day 6, and continued to decrease thereafter. Overall, we propose a unified multi-state approach for modeling infectious and chronic disease progression, which may provide evidence to inform timely and effective infectious disease prevention.

## 1. INTRODUCTION

Infectious diseases and chronic diseases are two major fields in epidemiology that have traditionally been studied separately because of their distinct etiologies. Infectious diseases are caused by transmissible pathogens, and disease transmission depends largely on the number of infected individuals in the population and the biological characteristics of the pathogen, rather than on individual-level factors.^1^ In contrast, the development and progression of chronic diseases are largely determined by individual-level factors. For example, in cardiovascular disease, major risk factors include body mass index, blood pressure, smoking status, and blood lipid levels, while disease progression are strongly affected by treatments such as lipid-lowering medication use and heart bypass surgery.^2^ These factors vary across individuals, leading to substantial between-person difference in the development and progression of cardiovascular disease.

Given the differences in disease etiologies, the methods used to model disease dynamics also differ. As infectious disease data are typically collected as population-level aggregated data, they are modeled using compartmental models. The most widely used is the susceptible– infectious–recovered (SIR) model,^3^ which divides the population into three compartments, susceptible (S), infectious (I), and recovered (R). While for chronic disease, the data are usually collected as individual-level survival data and analyzed using multi-state models.^4–6^ However, despite these differences, the progression of both infectious and chronic diseases can be considered as stochastic processes and therefore modeled within a unified framework. Previous studies have pointed out that the aggregated infectious disease data can be reconstructed into individual-level data,^7–10^ with survival models applied to estimate contact rate. However, these studies have largely focused on the transition from the S to I state, and few studies have simultaneously modeled the three-state transition from S to I to R.

In this paper, we use a discrete-time multi-state framework initially proposed in modeling chronic disease^6^ to model the three-state transition of infectious disease, and show how it works under two scenarios: 1) all S, I, and R states are observed, and 2) only the I state is observed, with the R state treated as latent. We first introduce and compare the underlying methodological foundations for modeling infectious disease and chronic disease dynamics, then show the link between compartment models and multi-state models, and finally present how the multi-state framework can be applied to model infectious disease progression under these two scenarios.

## 2. Compartmental models for infectious disease dynamics

Many infectious diseases, such as influenza, COVID-19, and measles, are characterized by short-term outbreaks. The transmission of these diseases has several features: 1) the disease process has a relatively short period, 2) transmission rate from S to I state is primarily determined by the proportion of infected individuals in the population rather than by individual-level risk factors, and 3) the data are typically collected from surveillance systems (i.e., population-level aggregated data). The SIR compartmental models are widely used to describe the dynamics of infectious disease transmission, which divides the disease in S, I, R states (**Figure 1a**).

**Figure 1.**
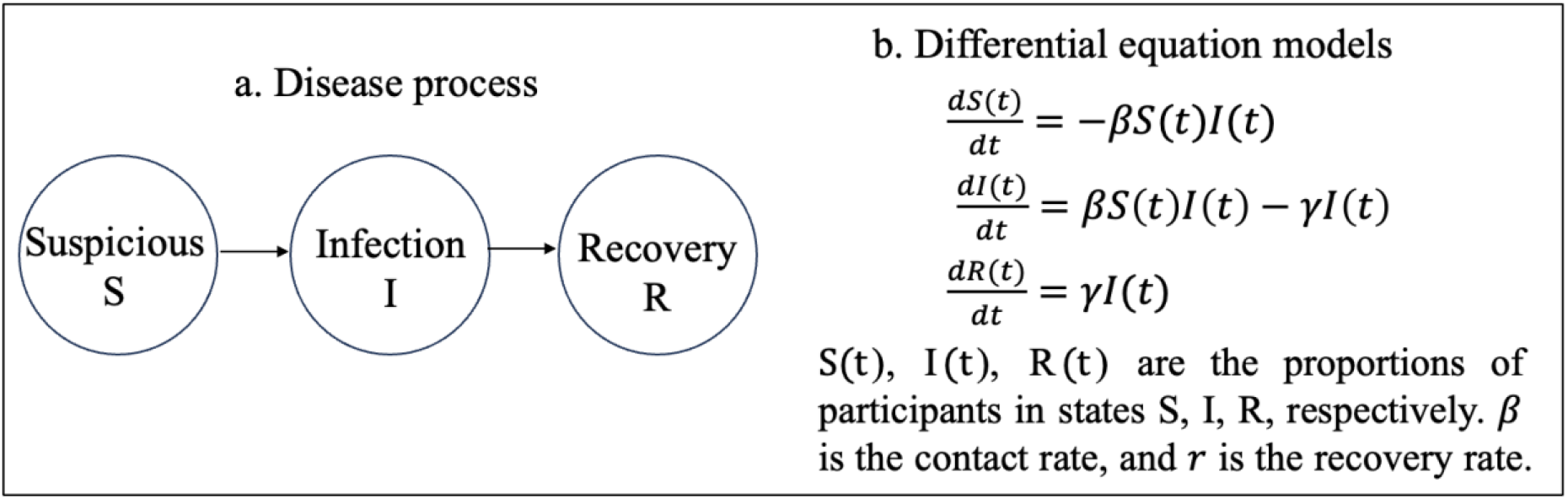
Differential equation models of infectious disease progression assuming a susceptible–infectious–recovered (SIR) process.

The changes in being in the three compartments over time, noted as *S*(*t*), *I*(*t*), and *R*(*t*), can be characterized using differential equations (**Figure 1b**). The equations estimate two parameters: 1) the contact rate, *β*, defined as the probability of transmitting the pathogen per contact; and 2) the recovery rate, *r*, defined as the rate at which infected individuals recover. Notably, *β* and *r* depend on pathogen-specific biological characteristics and are assumed to be the same across individuals. The transmission rate from S to I state is modeled as a function of the contact rate, *β*, and the proportion of individuals who are infected, i.e., *dS*(*t*)/*dt* = −*βS*(*t*)*I*(*t*). *β* and *r* determines the basic reproductive number (R_0_), defined as *β*/*r*. which indicates whether an outbreak will grow in its early stages. The value of R_0_ reflects the transmissibility of the disease and has public health implications for early prevention.

Parameters of compartment models can be estimated using method of moments, maximum likelihood estimation (MLE), or Bayesian estimation.^11, 12^ All methods use using the aggregated infectious data and model the probabilities of being in each state in discrete time. Method of moments solves the differential equations by assuming *S*(*t*)=1, and thus may only be accurate during the early phase of disease outbreak.^11, 13^ The MLE estimates *β* and *r* by assuming the per time unit change in infectious cases and recovered cases follow Poisson distribution.^12^ The Bayesian estimation assigns a prior in estimating the likelihood of being in infectious and recovered states over time.^14^

From a multi-state survival perspective, infectious disease dynamics can be viewed as a stochastic process with the S, I, and R states. The state occupancy probabilities (or survival probabilities) — defined as the proportion of participants in each state at time *t* — are the same as probabilities in each compartment, namely *S*(*t*), *I*(*t*), and *R*(*t*). The transition rate (or hazard) from the S to I state is defined as the instantaneous risk of infection among susceptible individuals at time *t*, which is estimated as *λ*_*SI*_ (*t*)*= βI*(*t*)*S*(*t*)/*S*(*t*)*=βI*(*t*). The transition rate from the I to R state is defined as the instantaneous recovery risk among infected individuals at time *t*, estimated as *λ*_*IR*_ (*t*)*= rI*(*t*)/*I*(*t*)*= r*. Notably, *λ*_*SI*_ (*t*) changes with time, as it depends on the proportion of infected participants at *t*, whereas *λ*_*IR*_ (*t*) is the same as recovery rate and constant over time.

## 3. Multi-state models for chronic disease dynamics

In contrast to infectious diseases, chronic diseases, such as cardiovascular disease, have a long course that may take decades to develop. The progression of chronic disease has three characteristics: 1) a long-term disease process, 2) strong effects of individual-level factors (such as lifestyle) on disease progression, and 3) data that are typically collected longitudinally in cohorts followed for decades, with risk factors measured at the participant level. We assume a three-state chronic disease progression, denoted as *S*_0_, *S*_1_, and *S*_2_ (**Figure 2a**). To be comparable to infectious disease, we assume *S*_0_ affects only *S*_1_ and *S*_1_ affects only *S*_2_.

**Figure 2.**
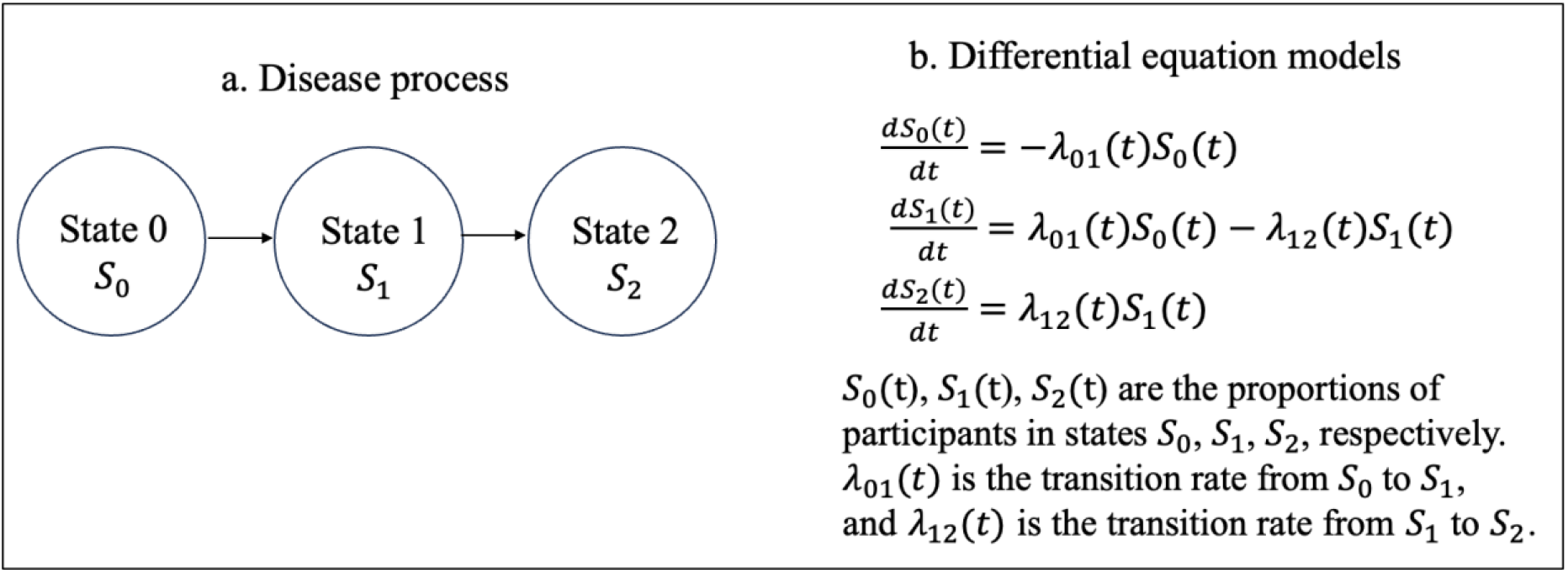
Differential equation models of chronic disease progression assuming a three-state process.

The differential equations for chronic disease progression can be written as **Figure 2b**. Like infectious disease models, these equations describe the instantaneous changes in the *S*_0_, *S*_1_, and *S*_2_ states over time; however, they are different in three aspects. First, the transition rate *λ*_01_ (*t*) is independent of the proportion of participants in *S*_0_ state, as chronic diseases are not transmissible. Second, *λ*_01_ (*t*) and *λ*_12_ (*t*) are largely determined by individual-level risk factors, whereas infectious disease models often assume homogeneous transition rates across individuals. Third, the transition rates *λ*_01_ (*t*) and *λ*_12_ (*t*) are time-varying, whereas in infectious disease models the transition rate *λ*_*SI*_ (*t*) is time-varying but *λ*_*IR*_ (*t*) is assumed to be constant. A comparison of infectious disease and chronic disease modeling is summarized in **Table 1**.

**Table 1.**
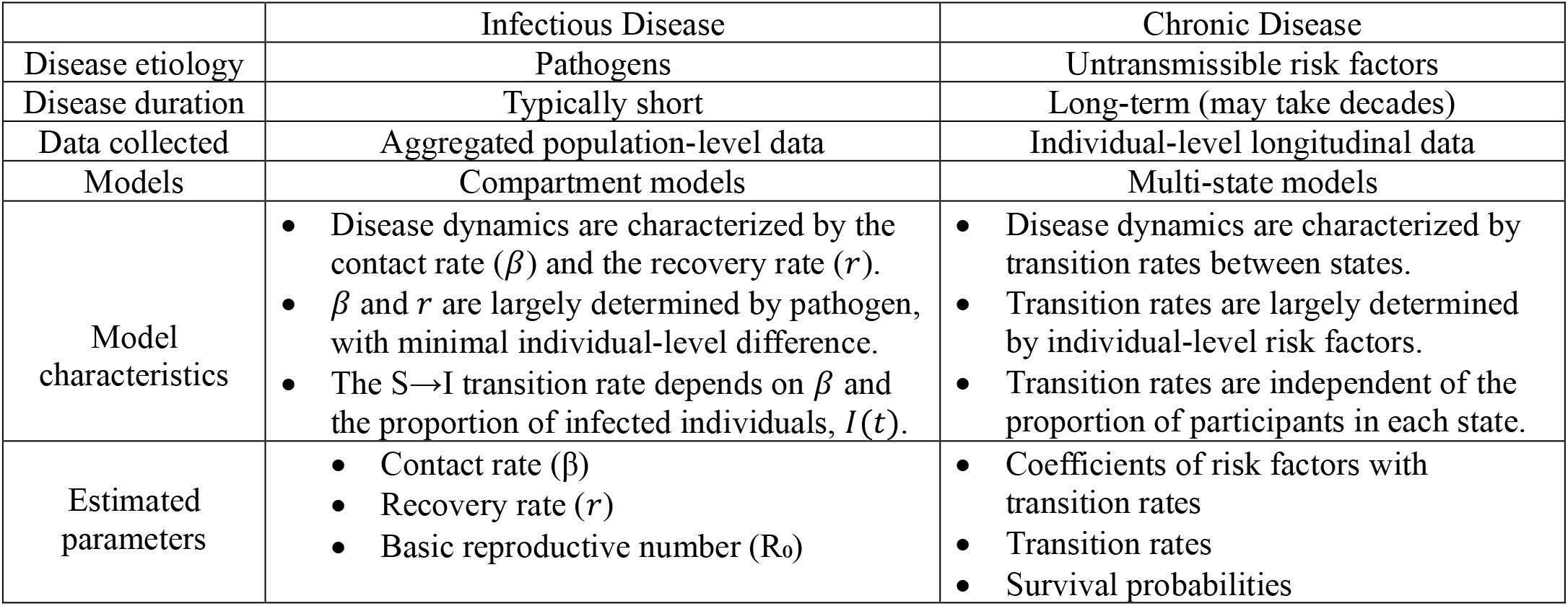
A comparison of infectious disease and chronic disease progression.

|  | Infectious Disease | Chronic Disease |
| --- | --- | --- |
| Disease etiology | Pathogens | Untransmissible risk factors |
| Disease duration | Typically short | Long-term (may take decades) |
| Data collected | Aggregated population-level data | Individual-level longitudinal data |
| Models | Compartment models | Multi-state models |
| Model characteristics | Disease dynamics are characterized by the contact rate ($\beta$) and the recovery rate ($r$).<br>$\beta$ and $r$ are largely determined by pathogen, with minimal individual-level difference.<br>The S→I transition rate depends on $\beta$ and the proportion of infected individuals, $I(t)$. | Disease dynamics are characterized by transition rates between states.<br>Transition rates are largely determined by individual-level risk factors.<br>Transition rates are independent of the proportion of participants in each state. |
| Estimated parameters | Contact rate ($\beta$)<br>Recovery rate ($r$)<br>Basic reproductive number ($R_0$) | Coefficients of risk factors with transition rates<br>Transition rates<br>Survival probabilities |

For parameter estimation of chronic disease, *λ*_01_ (*t*) and *λ*_12_ (*t*) are estimated using multi-state models,^4–6^ which is an extension of survival analysis from one disease endpoint to multiple endpoints. Basically, the models split a person’s follow-up time into subsets according to the disease state being in. Within each subset, the data structure is changed from wide to long format (Andersen-Gill format in survival analysis) by time (*e*.*g*., days, weeks, or years). Cox models (**Models 1, 2**) are fitted to each data subsets, with coefficients of *x*_1_,…, *x*_*m*_ indicating the associations of risk factors *x*_1_,…,*x*_*m*_ with transition rates.

**Model 1** in subset 1 starting from S_0_ state: *λ*_01_ (*τ, t,x*_1_,…, *x*_*m*_)= *λ*_01_ (*τ*)*exp*(*f* (*t*) + *x*_1_+…+*x*_*m*_),

**Model 2** in subset 2 starting from S_1_ state: *λ*_12_ (*τ, t,x*_1_,…, *x*_*m*_)= *λ*_12_ (*τ*)*exp*(*f* (*t*) + *x*_1_+…+*x*_*m*_), where *x*_1_,…, *x*_*m*_ are individual-level risk factors, *t* is the time interval in the long-data format, and is time to event in each time interval, and *λ*_01_ (*τ*) and *λ*_12_ (*τ*) are baseline hazards.

In this paper, we focus on a discrete-time multi-state framework due to three reasons.^6^ First, this framework estimates transition rates in discrete time by approximating rates by risks within a short time interval.^15^ This is different from multi-state Cox models which does not report transition rates (the reason is that Cox model is a semiparametric model where the baseline hazard is modeled non-parametrically and canceled out in the conditional likelihood estimation).^16^ Second, the estimation of transition rates in discrete time is suitable to infectious disease where the data are usually collected by days or weeks. Third, the discrete-time transition rates can be flexibly used to generate statistics with the 95% confidence interval estimated using bootstrap. These statistics include state occupation probability (or survival probabilities in each state) and the reproductive number, which are of public health significance in infectious disease epidemiology, as described in detail in **Section 4**.

## 4. Infectious disease progression is a special case of a multi-state process

Previous studies have pointed out the link between compartment models and survival analysis.^7–10^ The transition from the S to I state can be modeled by first reconstructing aggregated infection data into an individual-level population and then converting the data into a long-format structure to fit survival models. As the transition rate *λ*_*SR*_ (*t*) depends on *I*(*t*), *I*(*t*) can be included as a time-varying covariate in the model. However, these papers have primarily focused on the transition from *S*(*t*) to *I*(*t*), and few studies have simultaneously modeled the three-state process, *S*(*t*), *I*(*t*), and *R*(*t*). In this paper, we consider infectious disease progression as a special case of a multi-state process. We adopt the discrete-time multi-state framework to estimate the contact rate (*β*), recovery rate (*r*), and the basic reproductive number (*R*_0_), as well as transition rates and survival probabilities in each state over time. We also demonstrate how this framework can be used to estimate these parameters when only *I*(*t*) is observed.

### 4.1. SIR model with *S*(*t*), *I*(*t*), and *R*(*t*) observed

#### a. Calculation of the incident infected and recovery cases over time

Suppose in a closed population with *N* participants with *S*(*t*), *I*(*t*), and *R*(*t*) observed, where *t* is from *t*_1_ to *t*_*e*_ in discrete time. Based on this aggregated data, we can calculate the cases of being in each state over time, *N*_*s*_ (*t*), *N*_*I*_ (*t*), *N*_*R*_ (*t*). The incident recovered cases at time *t, N*_*IR*_ (*t*), and the incident infected cases, *N*_*SI*_ (*t*), can be calculated as shown in **Table 2**.

**Table 2.** Calculation of the incident infected and recovery cases over time.

|  | Data observed |  |  | Calculated cases |  |
| --- | --- | --- | --- | --- | --- |
| | Suspicious participants, $N_S(t)$ | Infection cases, $N_I(t)$ | Recovered cases, $N_R(t)$ | Incident recovery cases, $N_{IR}(t)$ | Incident infection cases, $N_{SI}(t)$ |
| $t_1$ | $S(t_1) \times N$ | $I(t_1) \times N$ | $R(t_1) \times N$ | $N_R(t_1)$ | $N_I(t_1)$ |
| $t_2$ | $S(t_2) \times N$ | $I(t_2) \times N$ | $R(t_2) \times N$ | $N_R(t_2) - N_R(t_1)$ | $N_I(t_2) - N_I(t_1) + N_{IR}(t_1)$ |
|  | .... | .... | .... | .... | .... |
| $t$ | $S(t) \times N$ | $I(t) \times N$ | $R(t) \times N$ | $N_R(t) - N_R(t-1)$ | $N_I(t) - N_I(t-1) + N_{IR}(t-1)$ |

#### b. Construction of the individual-level multi-state process

Given that there are no individual-level covariates, we construct a pseudo population with *N* participants, and randomly assign *N*_*SI*_ (*t*_1_) participants transitioned from S to I state at time *t*_1_. Then among the remaining *N*_*SI*_ (*t*_2_) susceptible participants, we randomly assign *N*_*SI*_ *(t*_2_)participants transitioned from S to I state at time *t*_2_, and so on until the end of follow up. Then among the participants who are infected cases at time t (the number of infection cases is equal to *N*_*I*_ (*t*)), we randomly select *N*_*IR*_ (*t*) as recovered cases. We do the random assignment at each time point until the end of follow up.

#### c. Creation of multi-state data subsets and change to long-format data structure

As described in detail in the paper that introduced the discrete-time multi-state framework,^6^ we split each person’s follow-up time into two subsets by disease state. Subset 1 starts from S state and ends in developing I state or end of follow up. Subset 2 starts from I state and ends in developing R state or end of follow up. Within each data subset, we discretize time into intervals by the unit the data are collected and change the data structure from wide to long form.

#### d. Estimation of contact rate 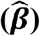 in data subset 1

Recall that in the compartment model in **Figure 1**, the incident infection cases are modeled as *N*_*SI*_ (*t*) / *N*_*S*_ (*t*) = *βI* (*t*) Given the long-format data structure, theoretically this could be modeled using a Cox model as **Model 1**. However, as infectious disease data are usually collected in discrete time, the time-to-event values within each time interval would be the same for everyone, and the Cox model would be reduced to a Poisson regression (**Model 3**).

**Model 3:** *log* (*λ*_*SI*_ (*t*)) =log *β*+1 × *log* (*I*(*t*)), where *λ*_*SI*_ (*t*) is the transition rate from S to I states and *λ*_*SI*_ (*t*) = *N*_*SI*_ (*t*_1_)/ *N*_*S*_(*t*_1_).

The coefficient of *log* (*I*(*t*)) is forced to be 1, given the assumption of linear transmission of the disease (The differential equation is *N*_*SI*_ (*t*)/ *N*_*S*_ (*t*) = *βI*(*t*)). Poisson model naturally assigns a variable with a coefficient of 1 as an offset, as reported in previous publications.^7–10^ The estimated intercept is 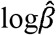, which is the contact rate in log scale.

In fact, the Poisson model allows for time-varying contact rate *β* (*t*) using **Model 4**, which assumes a *f*(*t*) function (such as linear, quadratic, spline function, etc) of contact rate in log scale with time. The corresponding compartment model equation is *N*_*SI*_ (*t*)/ *N*_*S*_ (*t*)= *β*(*t*) *I*(*t*).

**Model 4:** *log*(*λ*_*SI*_(*t*)) *=****K*** *f*(*t*) + *k*_0_ + 1 × *log* (*I*(*t*)),where ***K*** are the coefficients of *f*(*t*), *k*_0_ is the intercept, and 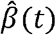 is estimated as 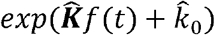.

The Poisson model can further allow for non-linear transmission of pathogen using **Model 5**, where the coefficient of *log* (*I* (*t*)) is not assigned as 1 but is truly estimated from the model. The corresponding compartment model equation is 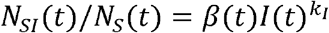.

**Model 5:** *log*(*λ*_*SI*_(*t*)) *=****K*** *f*(*t*) + *k*_0_ + *k*_1_ × *log* (*I*(*t*)), where 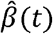 is estimated as 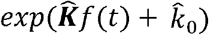.

We can use likelihood ratio tests to determine which model out of **Models 3-5** best fits the data.

#### e. Estimation of recovery rate 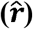 in data subset 2

Recall that the recovery rate is modeled as *N*_*IR*_ (*t*)/ *N*_*R*_ (*t*)=*r* for compartment model. For the long-format data, we model recovery rate (*r*) using a Poisson model without covariates into the model (**Model 6**). The estimated intercept would be 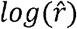.

**Model 6:** *log*(*λ*_*IR*_(*t*)) *= log* (*r*), where *λ*_*IR*_ (*t*) is the transition rate from I to R states and *λ*_*IR*_(*t*) = *N*_*IR*_ (*t*)/ *N*_*RI*_ (*t*)

The Poisson model also allows for time-varying recovery rate *r* (*t*) using **Model 7**. The corresponding compartment model equation is *N*_*IR*_ (*t*) / *N*_*R*_ (*t*) = *r*(*t*).

**Model 7:** *log*(*λ*_*SI*_(*t*)) *=****K***_***r***_ *f*(*t*) + *k*_*r*_, where ***K***_***r***_ are the coefficients of *f*(*t*),*k*_*r*_ is the intercept, and 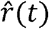 is estimated as 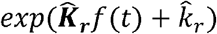.

We can use likelihood ratio tests to determine whether **Models 6 or 7** fits the data better.

#### f. Estimation of transition rate matrix, 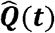, survival probability matrix, 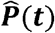 and reproductive number, 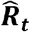

Under the estimated contact rate 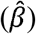 and recovery rate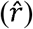, the transition rate from S to I state, 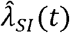, can be estimated from **Model 3**, and the transition rate from I to R states, 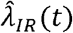, can be estimated from **Model 6**. The transition rate matrix is constructed as 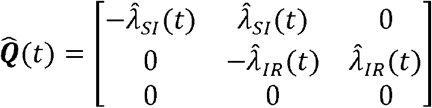. Using the Aalen-Johansen estimator (which is an extension of Kaplan-Meier estimator to a multi-state setting),^17^ the survival probability matrix can be estimated as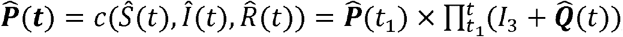, where *I*_3_ is an identity matrix. The reproductive number at time *t*,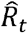, is defined as the ratio of those enter the infectious state over those recover from infectious state at time *t*, and can be estimated as 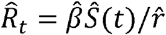 (or 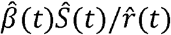 if *r* is time-varying). It accesses whether the number of participants in *I*(*t*) state will increase, with 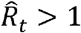 indicating that the disease is still expanding at time *t*.

The 95% confidence intervals of 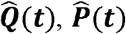 and 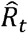 can be estimated using bootstrap. We randomly draw samples with replacement from the pseudo population created in Step b to generate a new sample that are the same size as the original population.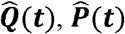, and 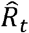 are estimated from each bootstrap sample. Statistically, the resulting parameter estimates across all bootstrap samples represent the empirical distribution and can be summarized with median values and a 95% confidence interval (CI) defined by 2.5th to 97.5th percentiles of the empirical distribution.

### 4.2. SIR model with only *I* (*t*) observed

In many infectious disease surveillance systems, due to the absence of routine follow-up to confirm recovery, only prevalent infected cases are collected, and information on the susceptible and recovered cases can only indirectly inferred. Our multi-state framework can deal with this scenario by considering recovery rate ( *r*) as a latent variable. We assume is constant over time and assign it a series of values between 0 and 1. Under each *r*, the incident infected and recovery cases over time can be calculated as a function of *r* (**Table 3**).

**Table 3.** Calculation of the incident infection and recovery cases over time only *I*(*t*) observed.

|  | Data observed | Data calculated |  |  |  |
| --- | --- | --- | --- | --- | --- |
| | Infection cases, $N_I(t)$ | Recovered cases, $N_R(t)$ | Suspicious participants, $N_S(t)$ | Incident recovery cases, $N_{IR}(t)$ | Incident infection cases, $N_{SI}(t)$ |
| $t_1$ | $I(t_1) \times N$ | $N_I(t_1) \times r$ | $N$ | $N_R(t_1)$ | $N_I(t_1)$ |
| $t_2$ | $I(t_2) \times N$ | $N_I(t_2) \times r$ | $N - N_I(t_1)$ | $N_R(t_2) - N_R(t_1)$ | $N_I(t_2) - N_I(t_1) + N_{IR}(t_1)$ |
|  | .... | .... | .... | .... | .... |
| $t$ | $I(t) \times N$ | $N_I(t) \times r$ | $N - N_I(t-1) - \sum_{i=1}^{t-1} N_R(t-1)$ | $N_R(t) - N_R(t-1)$ | $N_I(t) - N_I(t-1) + N_{IR}(t-1)$ |

Based Under *r* each, like addressed in **Section 4.1**, we construct a pseudo population, change the to long-format structure, and fit a Poisson model (**Model 3**) to estimate contact rate 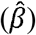. data on the *r* and 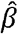, we can estimate the transition rate matrix, 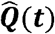, and the survival probability matrix, 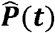. To identify the *r* that generates the estimates most close to the observed data, we calculate the sum of squared error (SSE) between observed and predicted infection cases under each *r*, and choose the the generate the smallest SSE, where 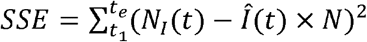.

## 5. APPLICATION

### 5.1. Study population

We used data from an outbreak of influenza in a British boarding school.^18^ In this closed population, 763 resident boys aged 10-18, experienced a sudden onset of influenza following the introduction of a single infectious individual in January 1978. Daily case counts were recorded over a two-week period, from January 22nd through February 4^th^, capturing the full course of the outbreak. The data were accessed via the R package “epimdr”,^19^ and are displayed in **Table 4**.

**Table 4.** Data of an influenza outbreak in a British boarding school in 1978.

| Day | Cases |
| --- | --- |
| 1 | 3 |
| 2 | 8 |
| 3 | 28 |
| 4 | 76 |
| 5 | 222 |
| 6 | 293 |
| 7 | 257 |
| 8 | 237 |
| 9 | 192 |
| 10 | 126 |
| 11 | 70 |
| 12 | 28 |
| 13 | 12 |
| 14 | 5 |

### 5.2. Data analysis

We assumed an SIR compartmental model of influenza transmission, with constant contact rate, constant recovery rate, and linear transmission of influenza from S to I state. Given that only infection cases, *N*_*I*_ (*t*), were known, according to **Table 3**, we estimated the recovered cases, *N*_*R*_ (*t*)= *N*_*I*_ (*t*) × *r*, the suspicious participants, 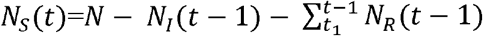, the incident recovery cases, *N*_*IR*_ (*t*) = *N*_*R*_ (*t*) − *N*_*R*_ (*t* −1), and incident infected cases *N*_*SI*_ (*t*) = *N*_*I*_ (*t*) − *N*_*I*_ (*t* −1). We chose *r* to be ranged from 0 to 0.48, which ensured that the total recovered cases ∑ _*t*_ *N*_*IR*_ (*t*) < *N* and the total infected cases ∑ _*t*_ *N*_*SI*_ (*t*) < *N*. We evaluated a series value of *r* between 0 and 0.48 at increments of 0.001. Under each value, we estimated the incident infected and recovery cases over time and constructed an individual-level pseudo population starting from S state and ending in I state or end of follow up. We format structure and fit a Poisson model (**Model 3**) to estimate contact rate 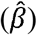, transition rate assumed 0 if the estimated incident cases had negative values. Then we changed the data to long-matrix, 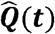 and survival probability matrix, 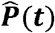. We chose the that minimized the sum of squared error between predicted and observed the infected cases, where 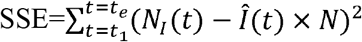.

### 5.3. Results

As shown in **Figure 3**, the estimated recovery rate 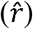 was 0.42, showing that 42% of the infected cases recovered each day. The corresponding contact rate 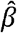 was 0.91 (95% CI:0.84, 0.98) (**Table 5**), and this high contact rate was due to that transmission occurred within a closed population. The basic reproductive number 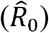 at day 1 was estimated to be 2.17 (95% CI: 2.00, 2.33), indicating that each infected person transmitted to on average 2.17 new cases. The transition rate from suspicious to infection increased rapidly from day 1, reached a peak around day 6, and declined afterwards due to the depletion of the susceptible population (**Figure 4**). The estimated probability of being in the infectious state followed a similar temporal pattern (**Figure 5**). The probability of being in the susceptible state decreased over time, whereas the probability of being in the recovery state increased. The reproductive number 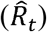 declined over the course of the outbreak, decreasing from 2.17 on day 1 to approximately 1 by day 6, and continued to decrease thereafter (**Figure 6**). Given that the number of infected cases increased before day 6 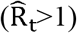 and decreased afterward 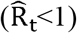, our finding showed that the turning point reached at around day 6, which provides key evidence to inform timely and effective infectious disease prevention.

**Table 5.** Estimated recovery rate (*γ*), contact rate (*β*), and baseline reproductive number (R_0_).

|  | Estimate<br>(95% confidence interval) |
| --- | --- |
| Recovery rate ($\gamma$) | 0.42 |
| Contact rate ($\beta$) | 0.91 (0.84, 0.98) |
| Baseline reproductive number ($R_0$) | 2.17 (2.00, 2.33) |

**Figure 3.**
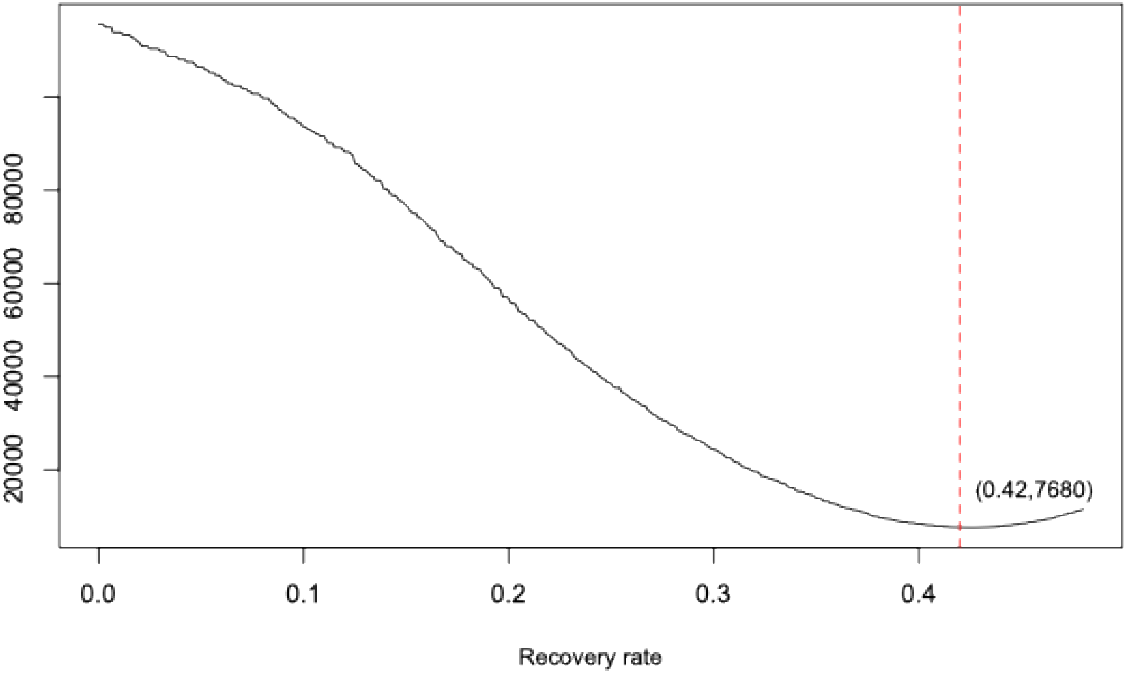
Sum of squared error in predicting infected cases across values of the recovery rate (*γ*).

**Figure 4.**
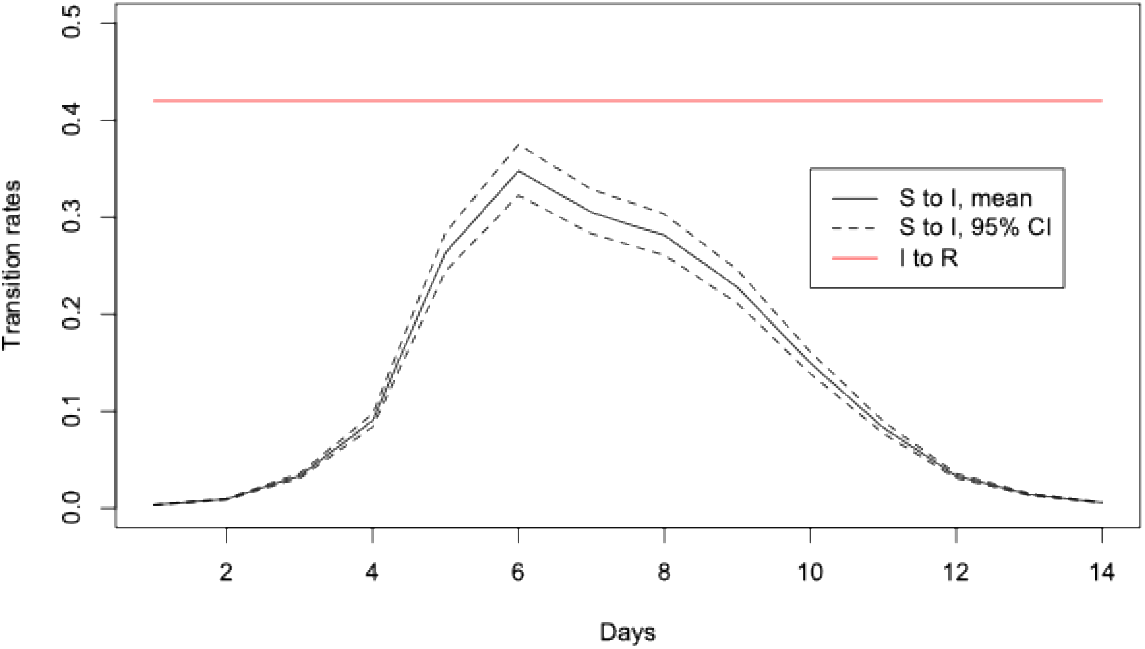
Estimated transition rates over time from susceptible (S) to infectious (I) and from infectious (I) to recover (R) states.

**Figure 5.**
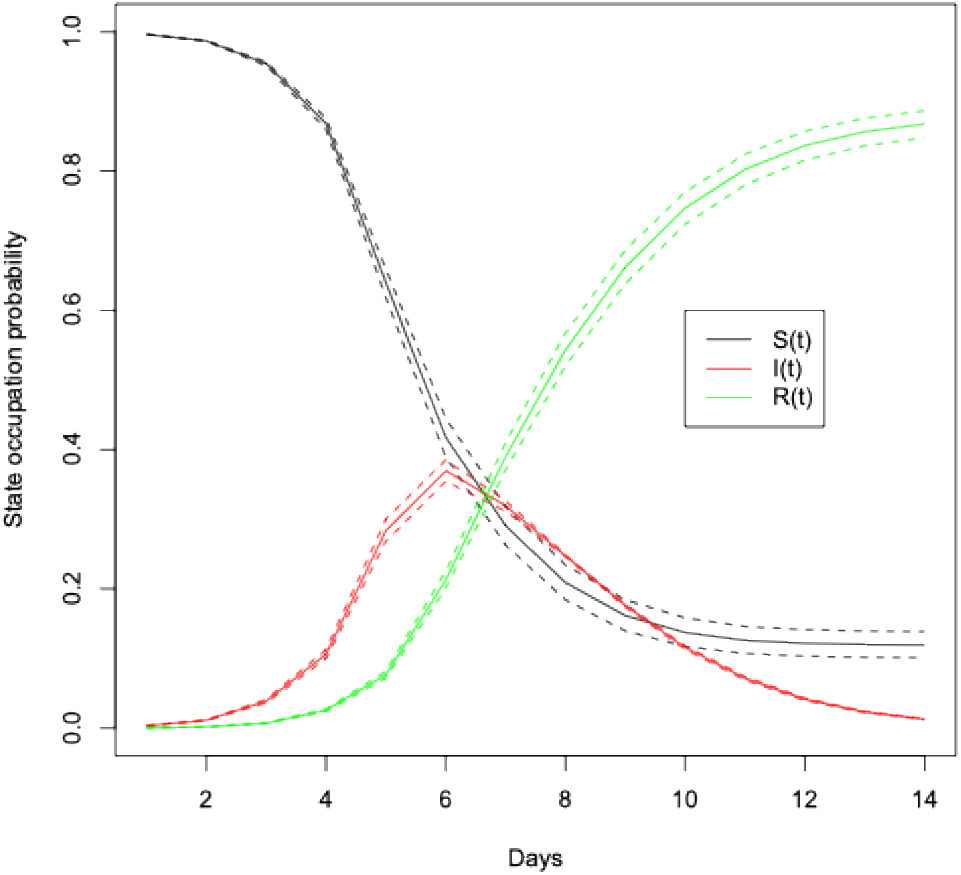
Estimated probabilities of being in the susceptible (S), infectious (I), and recover (R) states over time.

**Figure 6.**
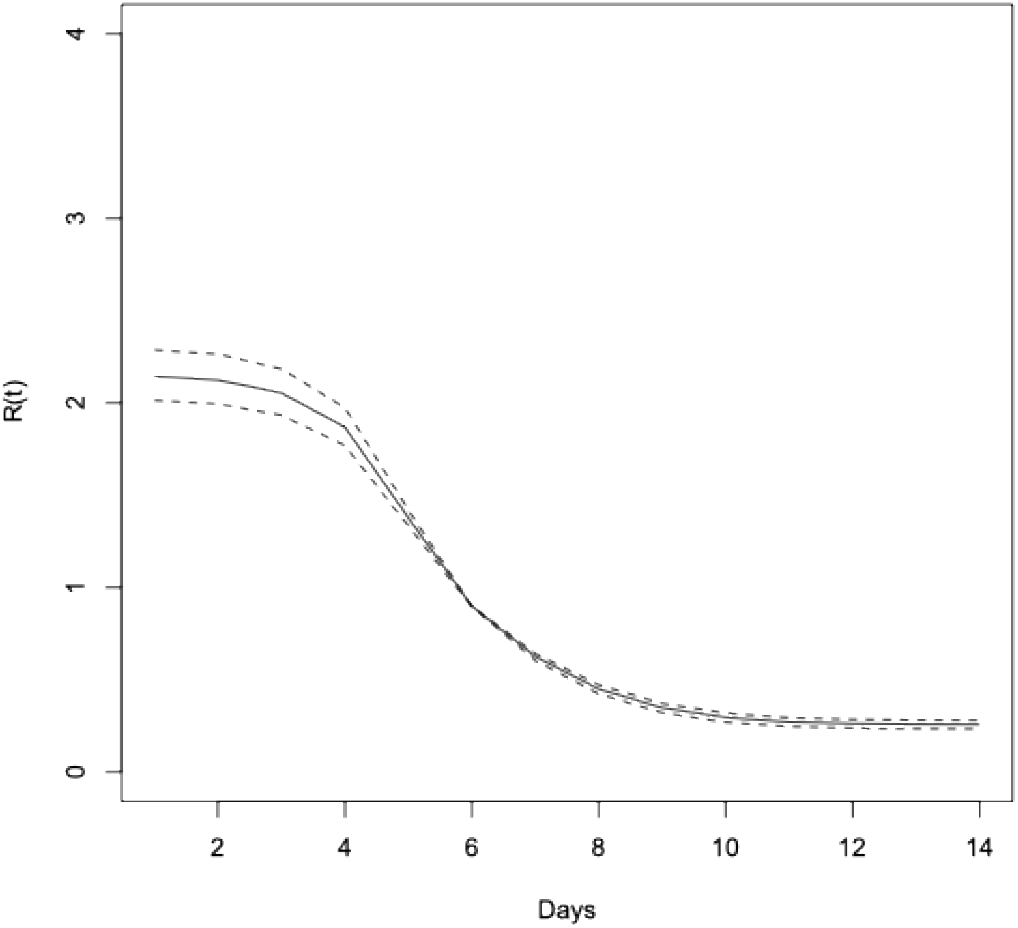
Estimated reproductive number (R_t_) over time.

## 6. DISCUSSION

In this paper, we use a discrete-time multi-state framework originally proposed for modeling chronic disease to model infectious disease progression. This can be achieved due to three reasons. First, as infectious disease progression does not depend on individual-level factors, the aggregated infectious data can be reconstructed into individual-level survival data. Second, to model that the proportion of infected people affects the transition rate from the S to I state, the proposed framework incorporates this proportion as a time-varying covariate into the Poisson model. Third, the discrete-time approach allows to flexibly generate statistics that is of high public health significance in infectious disease epidemiology, such as time-varying transition rates, survival probabilities, and reproductive number. The first two reasons have been pointed out in previous papers.^7–10^ However, we propose to adopt a multi-state approach to investigate the full three-states of infectious disease progression and explicitly showed how it works under scenarios with and without recovery state observed.

The methodological significance of our paper lies in its unification of infectious and chronic disease progression modeling. Traditionally, infectious and chronic disease have been two separate fields in epidemiology, each with its unique modeling methods. By examining their underlying methodological foundations, we demonstrate that both can be viewed as stochastic processes and can therefore be analyzed within a unified multi-state framework using individual-level data. In the future, it is of high interest to apply this multi-state framework to estimate parameters for Susceptible–Exposed–Infectious–Removed (SEIR) compartment model and to evaluate the intervention effect of vaccines.

With respect to the significance to infectious disease epidemiology, our approach estimates key parameters characterizing infectious disease transmission dynamics, such as contact rates, recovery rates, and reproductive numbers. Moreover, our framework enables the estimation of transition rates and state-specific survival probabilities over time, providing new evidence to infectious disease prevention. For example, in the influenza example, we showed that the transition rate from the suspicious to infection increased rapidly after day 1 and peaked around day 6. We also showed that the estimated probability of being in the infectious state followed a similar temporal pattern. Such information may help identify critical time windows for intervention and guide the timing of public health responses.

In summary, we propose a unified multi-state approach for investigating the progression of infectious and chronic diseases. By integrating concepts from multi-state survival analysis and infectious disease modeling, our approach may offer key evidence to inform timely and effective infectious disease prevention strategies.

## Data Availability

All data produced in the present study are available upon reasonable request to the authors

